# Anti-Platelet Factor 4 Antibody Clonal Heterogeneity and MGUS Status in HIT

**DOI:** 10.64898/2026.06.12.26355475

**Authors:** Adam J Kanack, Emily E Mauch, Mindy C. Kohlhagen, Jonathan Coker, David L. Murray, Anand Padmanabhan

**Affiliations:** Department of Laboratory Medicine and Pathology, Mayo Clinic, Phoenix, AZ; Department of Laboratory Medicine and Pathology, Mayo Clinic, Rochester, MN

## Abstract

**Background:** Monoclonal gammopathy of thrombotic significance (MGTS) is a recently described chronic prothrombotic condition characterized by monoclonal anti-PF4 antibodies that are detected above the polyclonal antibody background in patient sera (i.e. present as monoclonal gammopathy of undetermined significance, MGUS). Due to conflicting data in the published literature on antibody clonality in heparin-induced thrombocytopenia (HIT), we evaluated clonality and abundance of anti-PF4 antibodies in HIT, including investigating whether an MGUS, if present in HIT, represents the causative anti-PF4 antibody.

**Methods:** Blood samples from 15 patients with HIT were subject to Platelet Factor 4-dependent antigen-based and functional tests. The unmanipulated serum antibody repertoire and isolated anti-PF4 antibodies were subjected to mass spectrometric evaluation.

**Results:** Two of the 15 HIT patients had an IgG MGUS. Notably, anti-PF4 antibodies were not synonymous with the MGUS antibody in either of the two patients. Eight of the 15 patients demonstrated monoclonal anti-PF4 antibodies, however, none of the anti-PF4 antibodies were detectable as an MGUS upon evaluation of the entire serum antibody repertoire, reflecting their low abundance. In the seven patients with multiple anti-PF4 antibodies, non-monoclonality was confirmed by analysis of deglycosylated antibody heavy chains.

**Conclusions:** Anti-PF4 HIT antibodies are monoclonal in approximately 50% of HIT patients, however, antibody abundance is low such that they are not detectable over the polyclonal IgG background (i.e. are MGUS-negative), differentiating HIT from MGTS. This observation helps explain the transient nature of HIT relative to the persistent prothrombotic state seen in MGTS.

---

Heparin-induced thrombocytopenia (HIT) is a severe adverse effect of heparin characterized by production of platelet-activating anti-platelet factor 4 (PF4) antibodies which cause thrombocytopenia and thrombosis^1-3^. Recent developments in the anti-PF4 antibody field have revealed clonal restriction of anti-PF4 antibodies in vaccine-induced immune thrombotic thrombocytopenia (VITT)^4,5^ and in monoclonal gammopathy of thrombotic significance (MGTS)^6-11^. Studies to date in VITT, a primarily transient prothrombotic state, suggest that monoclonal antibodies are not detectable as a serum MGUS visible over the polyclonal antibody background, reflecting their low abundance^12^. In contrast, in MGTS, a serum detectable MGUS is the platelet-activating anti-PF4 antibody, accounting for the persistent nature of this thrombophilia^8,11^.

There is limited information on anti-PF4 antibody clonality in HIT. Previous studies have characterized HIT as a polyclonal antibody response based on the observation of non-overlapping epitopes revealed by cross-competition assays and experiments utilizing PF4 mutants and chimeras^13^, whereas a recent study proposes that HIT, like MGTS, is an MGUS-positive monoclonal anti-PF4 gammopathy^14^. Here we sought to examine three related but distinct questions in a cohort patients with HIT-1) Whether patients with HIT have a serum MGUS; 2) Whether a serum MGUS, if present, is synonymous with the anti-PF4 antibody, and 3) Whether anti-PF4 antibodies in HIT are mono- or polyclonal.

## METHODS

### Patients

Studies utilized remnant blood samples available after diagnostic testing was complete on 15 patients with HIT. Research studies were approved by the Institutional Review Board of Mayo Clinic.

### Anti-PF4 testing in Antigen-based and Functional assays

PF4 IgG testing (Immucor) was performed as ordered by the treating physician. Zymutest HIA IgG (Hyphen BioMed) immunoassay was performed according to manufacturer instructions. Serotonin release assay (SRA) testing was performed at a reference laboratory as ordered by the treating physician and the PF4-dependent P-selectin expression assay (PEA) was performed as previously described^15,16^ and as detailed in the *Supplementary Appendix*. Briefly, prostaglandin E1 was added to citrated whole blood obtained from healthy volunteers and centrifuged at 200 x g for 15 minutes to obtain platelet-rich plasma. Following an additional centrifugation step, platelets were resuspended in PBS/1.0% BSA and treated with PF4 (37.5 μg/mL) and patient sample. P-selectin expression was evaluated by flow cytometry.

### Anti-PF4 Antibody Isolation and deglycosylation

Anti-PF4 antibodies were isolated as previously described using PF4-treated heparin Sepharose beads^6,17^ and as detailed in the *Supplementary Appendix*. Briefly, heparin Sepharose beads were washed with PBS, pH 7.4 and incubated with PF4. Beads were then blocked with PBS/0.1% BSA. Following a thorough wash, patient samples were added to beads for 1 hour. After additional washes, eluates were obtained from the PF4/heparin Sepharose beads using 2M NaCl. Eluates were dialyzed against PBS before being evaluated in ELISA, PEA, and mass spectrometric studies. Antibody deglycosylation was performed with an N-glycosidase, PNGase F, per manufacturer instructions (New England Biolabs).

### Mass Spectrometry studies

“Mass-Fix” was performed on patient samples as previously^18,19^ described using a *Matrix-assisted laser desorption ionization time-of-flight Mass Spectrometry (*MALDI-TOF) mass spectrometer (Bruker Microflex LT, Germany) as detailed in the *Supplementary Appendix*. The basic method used for Liquid Chromatography Electrospray Ionization time-of-flight (LC-ESI-QTOF) mass spectrometry antibody analysis has been previously described.^20,21^ and as provided in the *Supplementary Appendix*. Antibodies were classified as polyclonal if more than one anti-PF4 antibody was identified.

## RESULTS

The fifteen patients comprised 11 males and 4 females with 14 European Americans and 1 African American. Mean patient age was 66 years and patients had 4Ts scores that were intermediate or high (median 5; range, 4-7; **Table1**). HIT serology was strongly consistent with HIT: The clinically used HIT ELISA in these patients (Immucor PF4 IgG) and serotonin release assay were positive in all patients (**Table 1**). An additional FDA-approved HIT ELISA (Zymutest HIA IgG), recently shown to have very high sensitivity for detection for pathogenic HIT antibodies^22^, was also positive in all patients (**Table 1**). Except for Patient 4, whose sample produced a borderline negative result in the PF4-dependent P-selectin expression assay (PEA, 15%; Positive cut-off 19%), all samples were also positive in that assay.

**Table 1.** HIT patient demographics, 4Ts score and serologic testing. Abbreviations: M – Monoclonal (8 patients); P – Polyclonal (7 patients); κ – *Kappa*; λ – *Lambda; EA-European American; AA-African American*.

| Patient<br>(Age-decade/Sex,<br>Race, 4Ts<br>Score) | Serum |  |  |  | Anti-PF4 Antibody |  |  |
| --- | --- | --- | --- | --- | --- | --- | --- |
|  | Immucor<br>PF4 IgG<br>(O.D.)<br>(Cutoff ≥<br>0.4) | Zymutest<br>HIA IgG<br>(O.D.)<br>(Cutoff ≥<br>0.3) | SRA<br>(%)<br>(Cutoff<br>≥ 20%) | PEA<br>(%)<br>(Cutoff<br>≥ 19%) | Zymutest<br>HIA IgG<br>(O.D.)<br>(Cutoff ≥<br>0.3) | PEA<br>(%)<br>(Cutoff<br>≥ 19%) | IgG Clonality<br>(Light Chain) |
| 1 (70sM,EA,5) | 2.32 | 3.27 | 100 | 82 | 3.10 | 108 | P (κ) |
| 2 (60sM,EA,7) | 2.25 | 3.08 | 94 | 84 | 2.90 | 107 | M (κ) |
| 3 (60sM,EA,4) | 1.50 | 2.29 | 83 | 56 | 1.40 | 102 | M (λ) |
| 4 (50sM,EA,6) | 1.72 | 1.69 | 92 | 15 | 0.99 | 91 | P (λ) |
| 5 (70sM,EA,5) | 1.37 | 2.57 | 100 | 77 | 1.42 | 101 | P (κ) |
| 6 (80sM,EA,5) | 1.28 | 3.53 | 97 | 99 | 3.06 | 99 | P (κ) |
| 7 (60sM,EA,5) | 1.84 | 2.37 | 90 | 92 | 0.71 | 13 | P (λ) |
| 8 (40sM,AA,7) | 1.19 | 1.96 | 63 | 92 | 0.63 | 96 | P (λ) |
| 9 (30sF,EA,7) | 1.40 | 2.77 | 99 | 52 | 1.74 | 103 | P (κ) |
| 10 (60sF,EA,7) | 1.42 | 3.95 | 83 | 126 | 3.67 | 108 | M (λ) |
| 11 (80sF,EA,4) | 2.07 | 3.61 | 97 | 103 | 3.41 | 107 | M (κ) |
| 12 (70sM,EA,5) | 1.66 | 2.67 | 92 | 56 | 1.82 | 100 | M (κ) |
| 13 (60sM,EA,4) | 1.47 | 2.9 | 97 | 82 | 1.92 | 115 | M (κ) |
| 14 (60sM,EA,6) | 1.18 | 1.84 | 94 | 76 | 0.31 | 101 | M (λ) |
| 15 (60sF,EA,6) | 1.44 | 2.19 | 65 | 99 | 2.38 | 99 | M (λ) |

Mass spectrometric antibody evaluation in the highly sensitive Mass-Fix diagnostic assay revealed an MGUS in only two patients with low monoclonal IgG concentrations of 40.5mg/dL and 31.4mg/dL (**Fig 1A-B**). Study of the isolated anti-PF4 antibodies revealed that in neither case was the MGUS synonymous with the anti-PF4 antibody (**Fig 1C-D**). Mass-Fix was negative for IgG MGUS in the 13 other patients of the cohort (**Supplementary Fig S1**).

**Figure 1.**
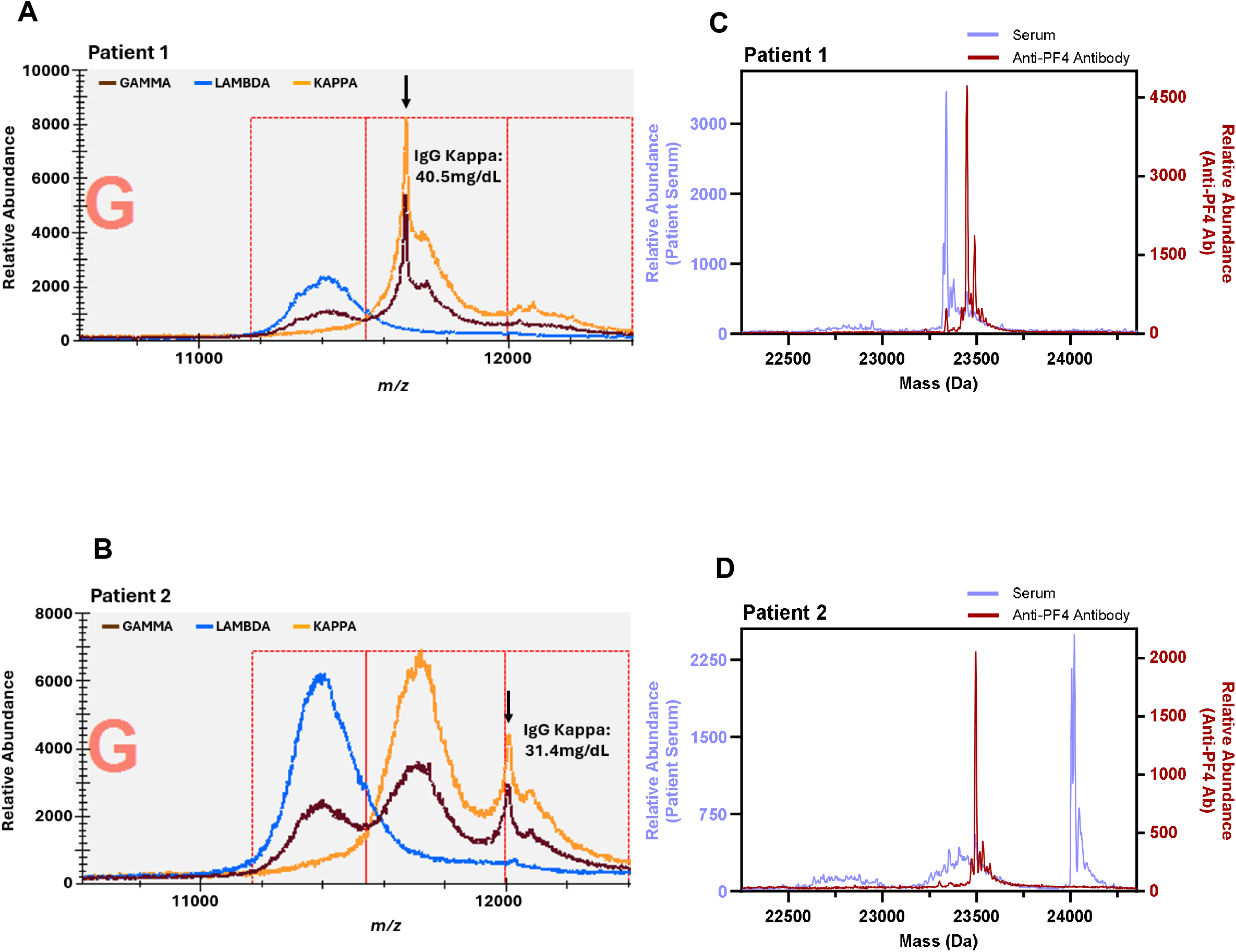
The serum MGUS was not an anti-PF4 antibody in the two MGUS-positive HIT patients. Panels A and B show Mass-Fix spectra of serum immunoglobulins from Patients 1 and 2. Brown traces denote light chains after IgG heavy chain (gamma) immunoenrichment, blue traces denote light chains after the immunoenrichment of lambda-containing immunoglobulins, and orange traces represent light chains after the immunoenrichment of kappa-containing immunoglobulins; arrows identify the monoclonal IgG kappa peaks, with concentrations indicated. Panels C and D show LC-ESI-QTOF calculated light-chain mass spectra of total serum IgG (blue) and affinity-purified anti-PF4 antibodies (red). In both patients, the isolated anti-PF4 antibody peak did not coincide with the serum MGUS peak, indicating that the MGUS was not synonymous with the HIT anti-PF4 antibody. PF4 denotes platelet factor 4, HIT heparin-induced thrombocytopenia, MGUS monoclonal gammopathy of undetermined significance, and LC-ESI-QTOF liquid chromatography electrospray ionization quadrupole time-of-flight mass spectrometry.

Successful affinity purification of anti-PF4 antibodies from patients was confirmed by antigen-based and functional HIT assays (**Table 1**). Patient 1’s anti-PF4 antibody appeared biclonal (**Fig 1C**), while Patient 2’s antibody was monoclonal (**Fig 1D**). Anti-PF4 antibodies from the remaining 13 patients demonstrated six more with polyclonal anti-PF4 antibodies (**Fig 2 & Table 1**), and seven more with monoclonal anti-PF4 antibodies (**Supplementary Fig S2 & Table 1**). Because glycosylation can broaden intact heavy-chain mass distributions and obscure clonality, PNGase F treatment was used to determine whether apparent polyclonal heavy-chain patterns persisted after removal of N-linked glycans. Among the seven non-monoclonal samples selected for deglycosylation, five had sufficient yield for heavy-chain analysis, and all five retained multiple heavy-chain peaks after PNGase F treatment (**Fig 3A-E**). Consistent with the monoclonal anti-PF4 light chain spectra seen for Patient 10 (**Supplementary Fig S2**), study of the patient’s anti-PF4 antibody deglycosylated heavy chain demonstrated a single clone (**Fig 3F**). As expected, deglycosylation of anti-PF4 antibodies did not shift light-chain profiles, consistent with the absence of appreciable N-glycosylation in light chains^23^ (**Supplementary Fig S3**). Mass/charge spectra of deglycosylated anti-PF4 antibody heavy- and light-chains are juxtaposed and presented in **Fig 4**, which demonstrates that multiple light chain and heavy chain peaks are seen in the polyclonal patient subset (**Fig 4A-E)**, and a single dominant light chain and heavy chain peak in the representative monoclonal anti-PF4 antibody sample from Patient 10 (**Fig 4F**).

**Figure 2.**
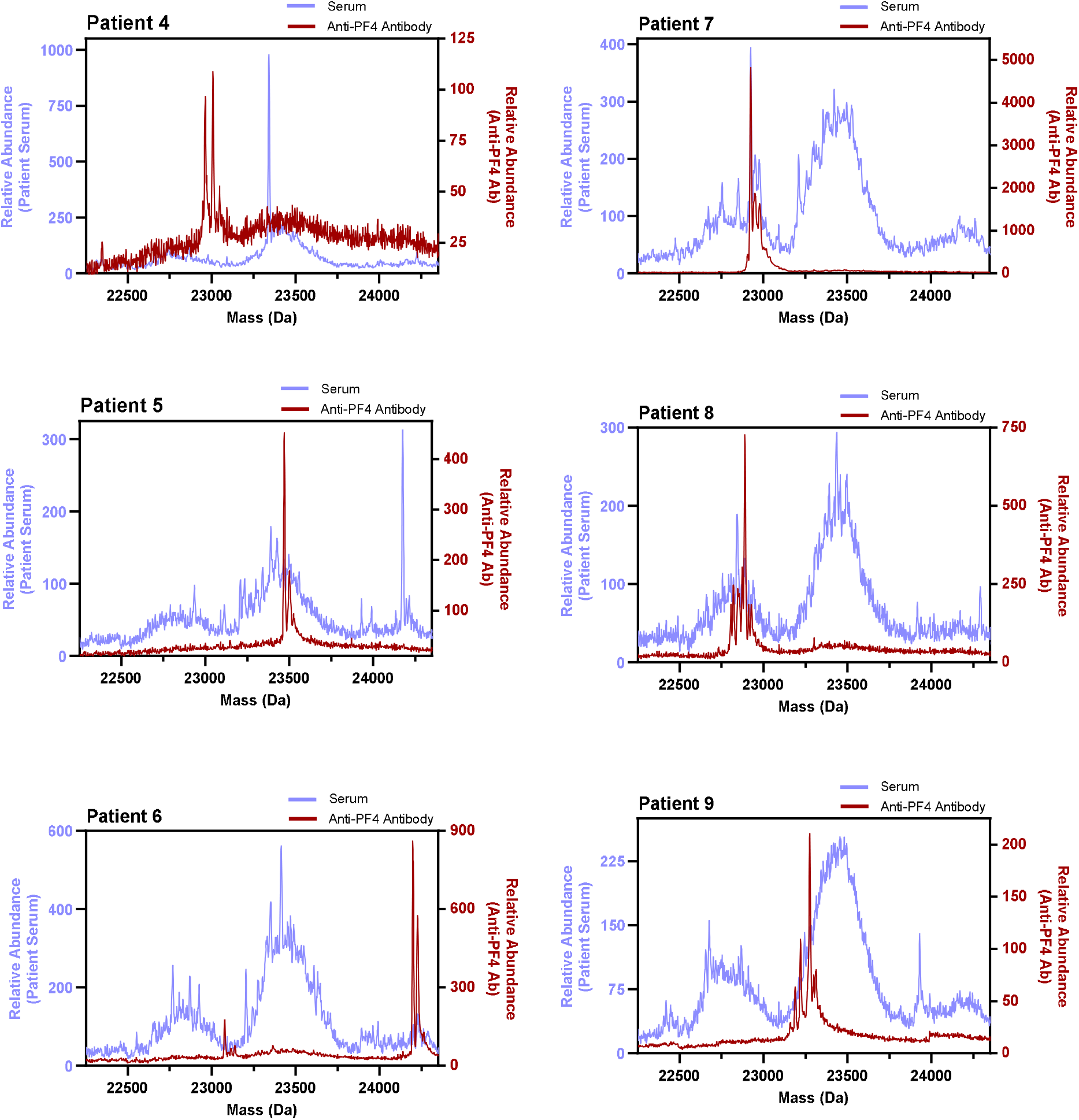
Polyclonal Anti-PF4 antibodies in HIT. LC-ESI-QTOF calculated light-chain mass spectra are shown for total serum IgG (blue) and affinity-purified anti-PF4 antibodies (red) from six HIT patients with polyclonal anti-PF4 antibodies, in addition to Patient 1 (Fig 1C). Multiple peaks within the purified anti-PF4 fraction indicate polyclonal anti-PF4 antibodies. Axes show calculated molecular mass and relative abundance. PF4 denotes platelet factor 4, HIT heparin-induced thrombocytopenia, and LC-ESI-QTOF liquid chromatography electrospray ionization quadrupole time-of-flight mass spectrometry.

**Figure 3.**
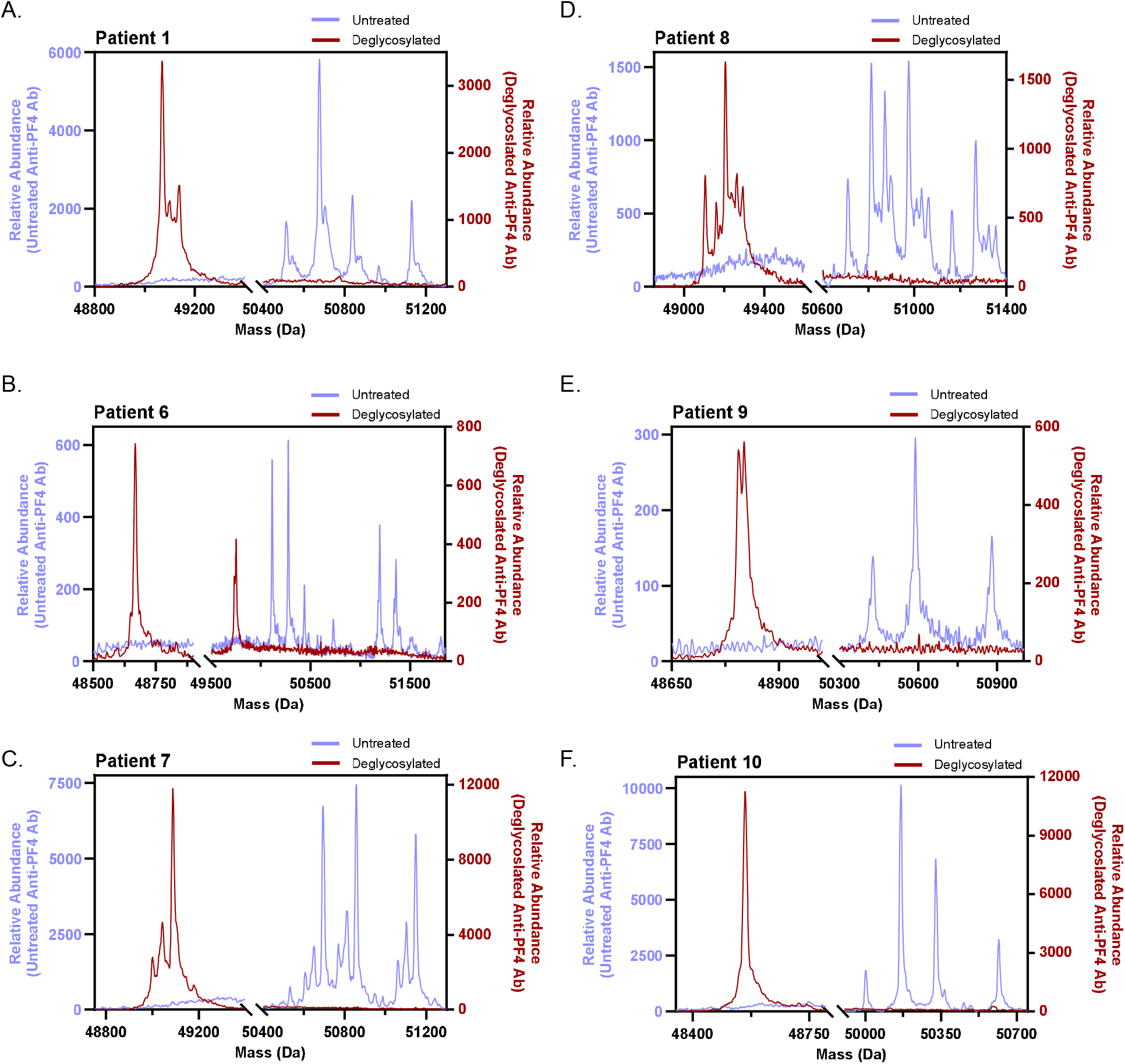
Deglycosylated heavy-chain spectra supports polyclonality of isolated anti-PF4 antibodies. Panels A through E show LC-ESI-QTOF calculated heavy-chain mass spectra of affinity-purified anti-PF4 antibodies from patients before (blue) and after PNGase F deglycosylation (red). Deglycosylation preserved multiple heavy-chain masses for Patients 1, 6, 7, 8, and 9, supporting a polyclonal anti-PF4 response in these patients. Panel F shows a representative monoclonal anti-PF4 antibody (Patient 10), in which deglycosylation yielded a single dominant heavy-chain mass, suggesting the production of a monoclonal anti-PF4 antibody in this patient. PNGase F denotes peptide:N-glycosidase F.

**Figure 4.**
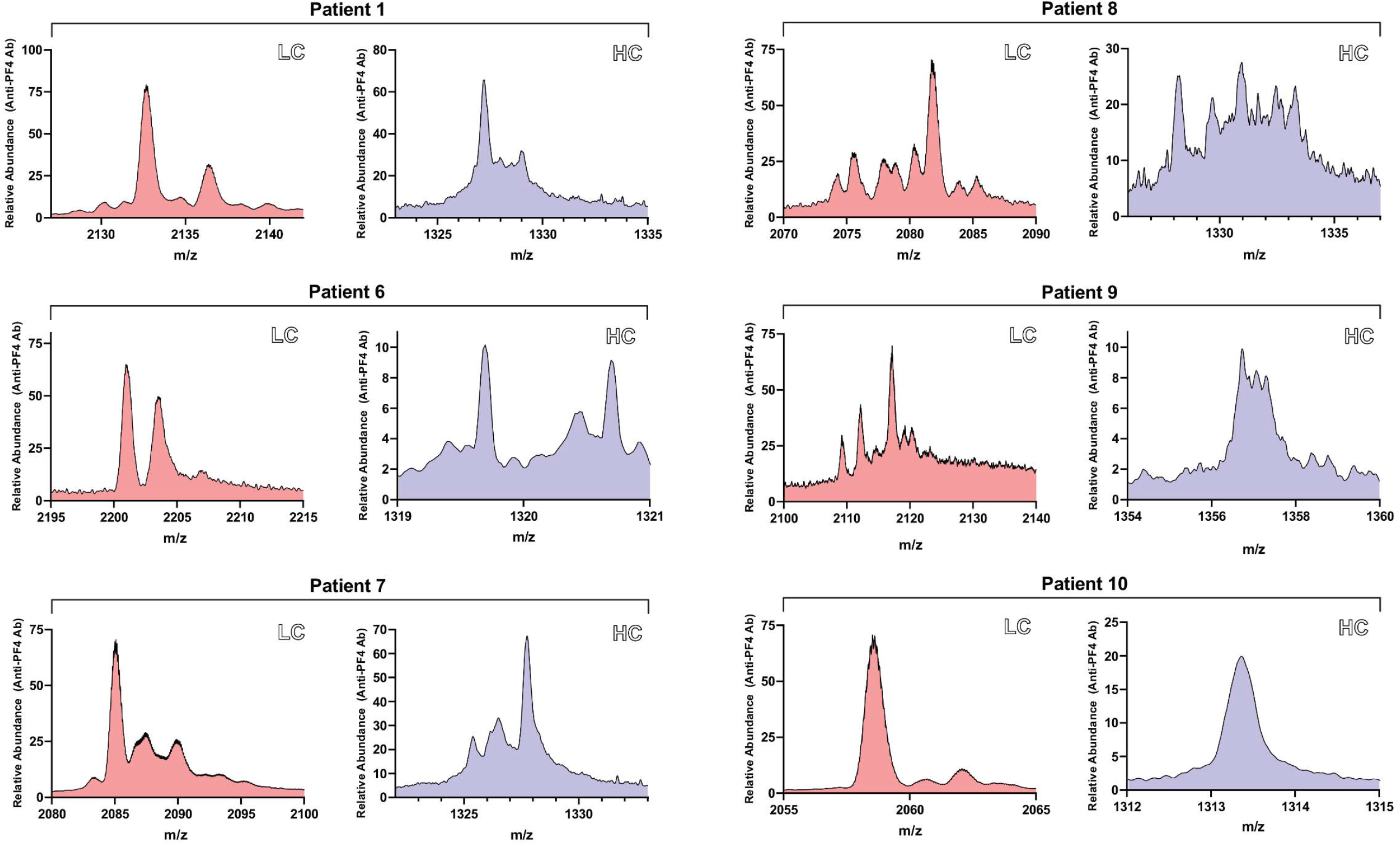
High-resolution paired light- and heavy-chain spectra after deglycosylation. Shown are high-resolution LC-ESI-QTOF mass/charge spectra of deglycosylated light (LC; orange) and heavy chains (HC; purple) from affinity-purified anti-PF4 antibodies. Panels A through E show patients with multiple light- and heavy-chain peaks, consistent with polyclonal anti-PF4 antibodies. Panel F shows Patient 10, a representative monoclonal anti-PF4 antibody, with a concordant single dominant light-chain and heavy-chain peak. LC denotes light chain, HC heavy chain, PF4 platelet factor 4, and LC-ESI-QTOF liquid chromatography electrospray ionization quadrupole time-of-flight mass spectrometry.

## DISCUSSION

MGUS is a persistent premalignant plasma-cell proliferative disorder^24^ that can progress to plasma cell cancers, and in some patients, causes non-malignant complications referred to as monoclonal gammopathy of clinical significance (MGCS). One recently identified sub-entity of MGCS is MGTS^10^ in which a monoclonal anti-PF4 MGUS antibody causes a chronic prothrombotic state due to the persistent nature of MGUS antibodies. In this context, HIT has been historically viewed as a polyclonal antibody-mediated disorder^13,25-27^. In recent work, *Treverton et al*.^14^ suggest that HIT is primarily an MGUS-positive state, with an MGUS detected in a majority of patients studied in their report (6/9, 67%). However, permanence associated with MGUS is inconsistent with the natural history of HIT, a transient prothrombotic state as noted in a recent comment in the *journal* to *Treverton et al*^28^.

Understanding HIT antibody clonality has important implications beyond understanding disease pathogenesis, including developing improved diagnostics and therapeutic interventions. This study separates three questions that have sometimes been conflated: whether patients with HIT have an incidental serum MGUS, whether the serum MGUS is synonymous with the anti-PF4 antibody, and whether the antigen-enriched anti-PF4 response is monoclonal or polyclonal. Using a highly sensitive mass spectrometry diagnostic test (Mass-Fix), IgG MGUS was seen in only 13% (2/15) of HIT patients, not significantly different from the reported IgG MGUS prevalence using this technique (7% in European Americans and 10% in African Americans older than 50 years of age)^29^. Notably, this prevalence was significantly lower than that reported recently (67%)^14^. The high sensitivity of the Mass-Fix technique (with a positive cut-off level of 20mg/dL for antibody detection) argues against lack of sensitivity of this technique as a reason for the divergent interpretation. In fact, prior work has demonstrated that MGUS estimates using MASS-FIX were at least threefold higher than prevalence estimates using conventional methods alone (serum protein electrophoresis [SPEP] followed by immunofixation electrophoresis [IFE])^29^.

Similar to the study of *Treverton et al*^*14*^., patients in our HIT cohort had convincing clinical and laboratory evidence of HIT with HIT ELISA optical densities >1.0, median 4Ts scores of 5 and positive SRAs. The causes of divergence between studies are unclear, however, some possibilities are: 1) The representative IFE gels presented in their study do not appear to clearly demonstrate a restricted monoclonal band and faint bands in IFEs which were listed as a “non-quantifiable” amounts of <1g/dL^14^ can cause significant inter-observer variability in interpretation, increasing the risk of false-positive calling^30^. 2) While Treverton et al., used mass spectrometry to study isolated anti-PF4 antibodies, it was not performed/presented on the total IgG repertoire to confirm the serum IFE findings that formed the basis of their conclusion that most HIT patients were positive for an MGUS, and, 3) In the Treverton et al., study, the disappearance of the poorly resolved IFE bands upon anti-PF4 depletion was used to conclude that the anti-PF4 antibody and MGUS were synonymous. In contrast, the mass spectrometry used in our study allowed for “fingerprinting” the antibody by mass/charge, and the data clearly demonstrate that anti-PF4 antibodies were not identical to the MGUS in the two MGUS-positive HIT patients in our cohort. The limited cohort size of our study does not preclude the possibility that rare HIT patients may develop high titer antibodies that may present as an MGUS, however, our findings suggest that it would be the exception rather than the rule. Partially consistent with the findings of *Treverton et al*, we found that eight HIT patients (∼50%) had monoclonal anti-PF4 antibodies, and the remainder (seven) had polyclonal antibodies.

Deglycosylated heavy-chain analysis further supported the presence of polyclonal anti-PF4 antibodies in the evaluable non-monoclonal cases.

In patients with polyclonal anti-PF4 antibodies, the multiple antibody peaks observed were not distributed across the entire mass range. The small mass differences between these species are consistent with the possibility that some HIT patients develop clonally related anti-PF4 antibodies that have diversified through somatic hypermutation following initiation of the immune response. Overall, these results are consistent with recent work showing that another transient anti-PF4 disorder, vaccine-induced immune thrombotic thrombocytopenia (VITT) is an MGUS-negative state^12^. The transient nature of HIT aligns with our findings of MGUS negativity, even though HIT patients may produce monoclonal antibodies in a subset of patients. These findings distinguish HIT from MGTS, in which the pathogenic anti-PF4 antibody is a high abundance overt serum monoclonal gammopathy. Implications of these findings on diagnostic and therapeutic development in these disorders should be considered.

## Data Availability

All data produced in the present study are available upon reasonable request to the corresponding author.

## AUTHOR CONTRIBUTIONS

A.J.K. and E.E.M. performed the laboratory studies and provided technical/scientific input.

M.C.K. provided critical technical support for mass spectrometry (MS) studies. J.C. assisted with MS analysis. D.L.M. provided scientific input related to MS studies. A.P. conceived the experimental plan and directed the laboratory studies. A.J.K., E.E.M, and A.P. wrote the first draft; all authors provided input and approved the final version.

## ACKNOWLEDGMENTS

We thank Stephanie Hafner and Jill Kappers from Mayo Clinic’s Research Innovation Office for their exceptional support in research coordination. This work was supported, in part, by National Institutes of Health grants HL158932 (AP) and HL171911 (AJK).

