## Supplementary Appendix for "Anti-Platelet Factor 4 Antibody Clonal Heterogeneity and MGUS Status in HIT"

**Table of Contents**

**Figures…………………………………………………………………………………………………………….2**

**Detailed Methods.……………………………………………………………………………………………….5**

**References……………………………………………………….……………………………………………….8**

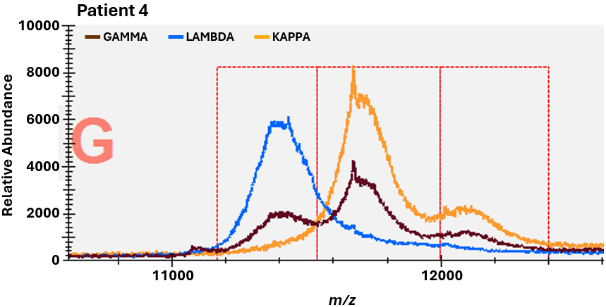

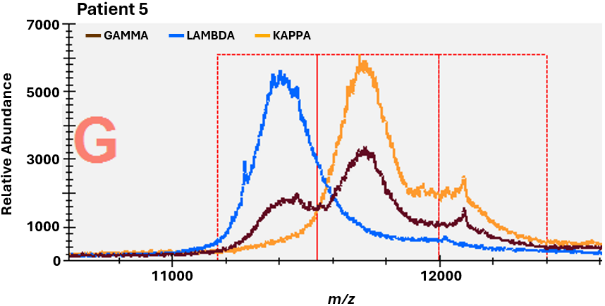

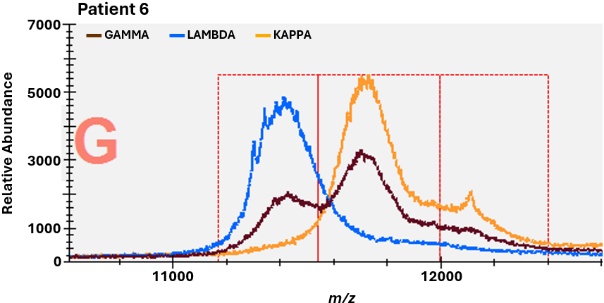

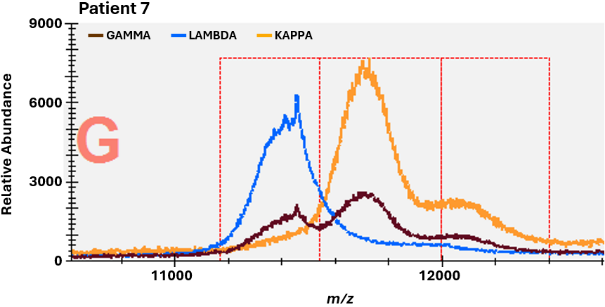

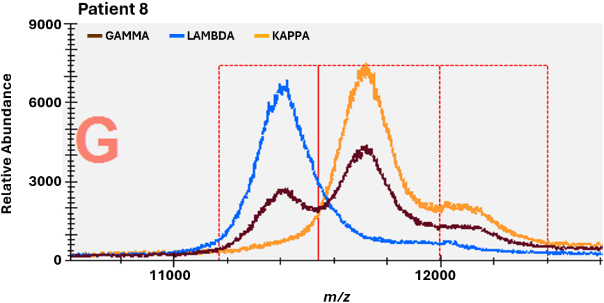

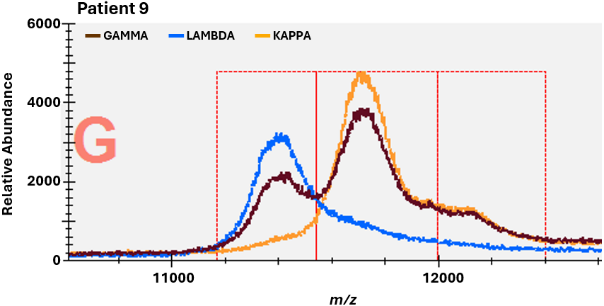

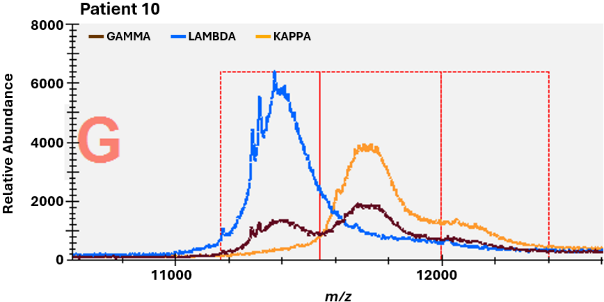

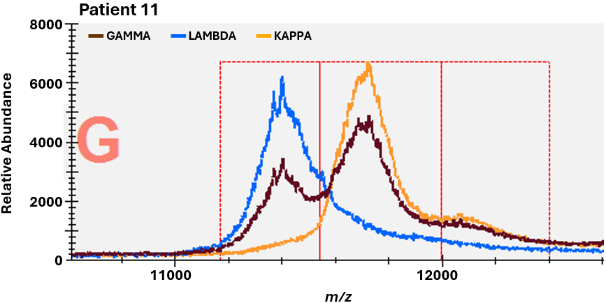

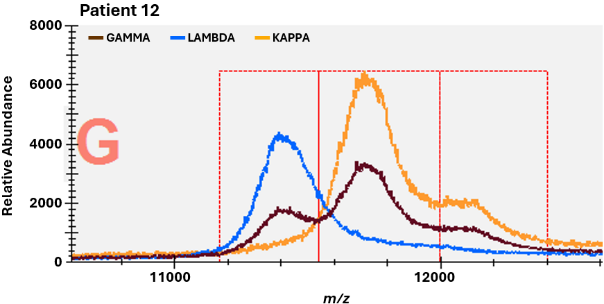

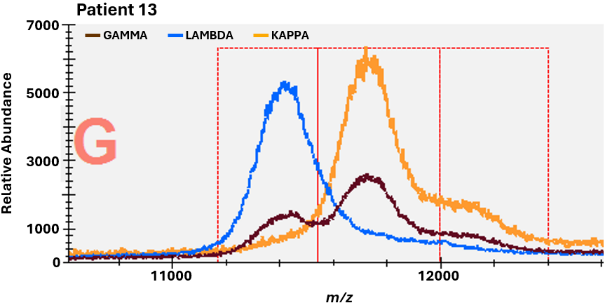

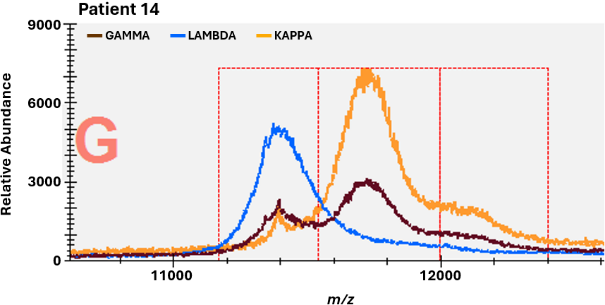

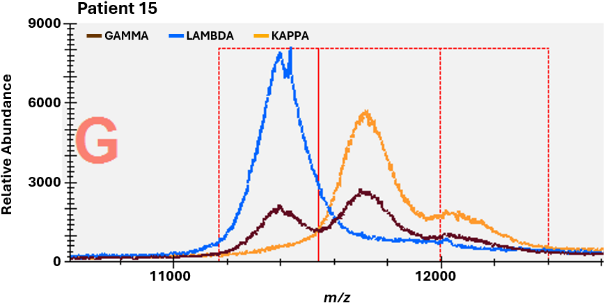

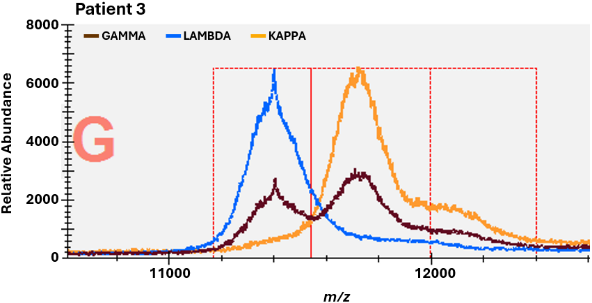

**Figure S1. IgG monoclonal gammopathies (MGUS) were not detected in 13 HIT patients.** Mass-Fix +2 spectra of patients 3-15 for IgG heavy chain (GAMMA) in brown, *lambda* containing Igs (LAMBDA) in blue, and *kappa* containing Igs (KAPPA) in orange. The x-axis depicts mass-charge (*m/z*), and the y-axis presents the relative abundance of identified antibodies. No monoclonal antibodies were detected (positive cut off: 20mg/dL )

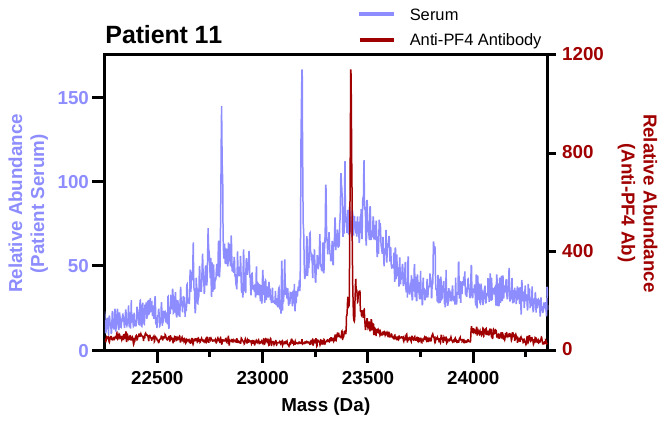

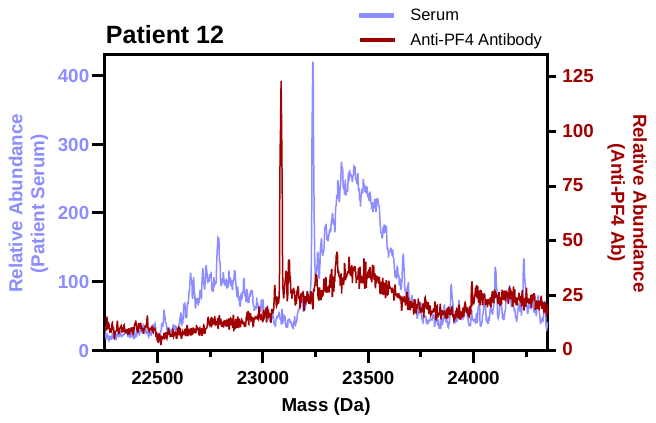

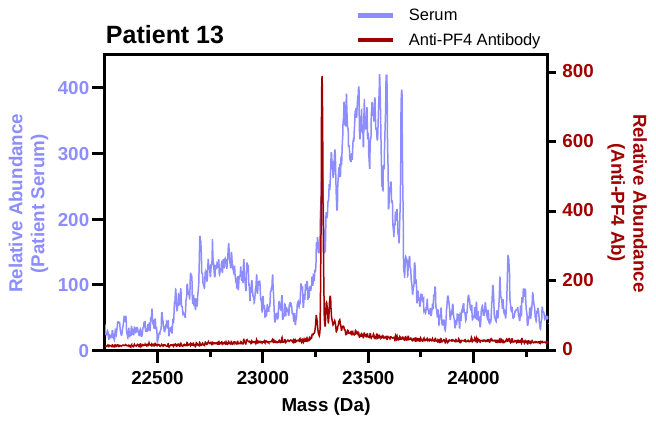

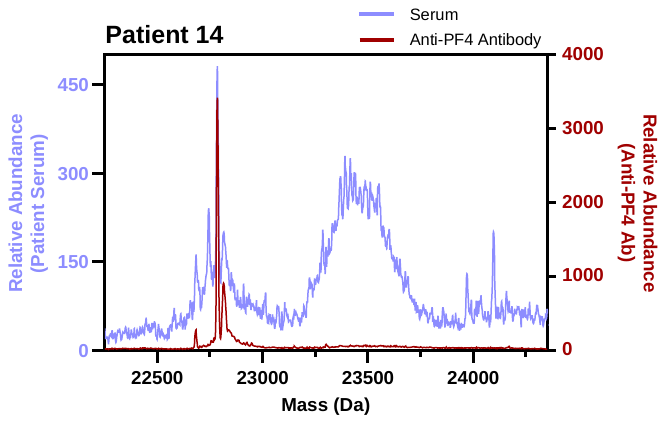

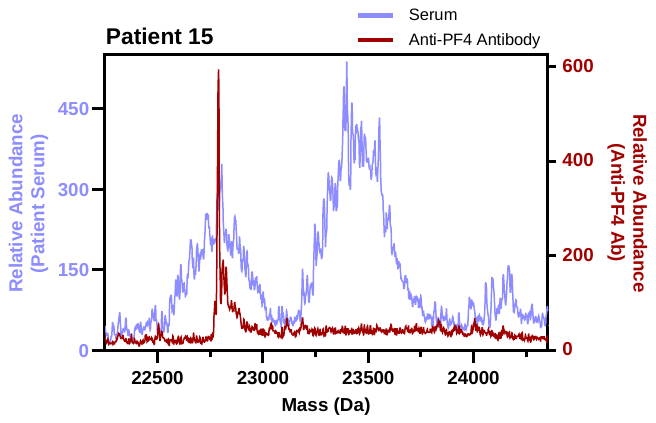

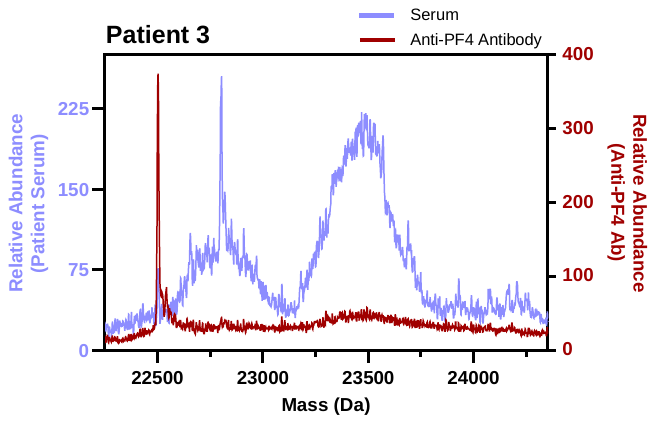

**Figure S2. HIT patients 3 and 10-15 display monoclonal anti-PF4 antibodies. (A-G)** LC-ESI-QTOF calculated mass spectra (Daltons) of light chain distributions associated with IgG heavy chains of patient serum in blue and isolated anti-PF4 antibodies in maroon. The y-axis presents the relative abundance of identified antibodies.

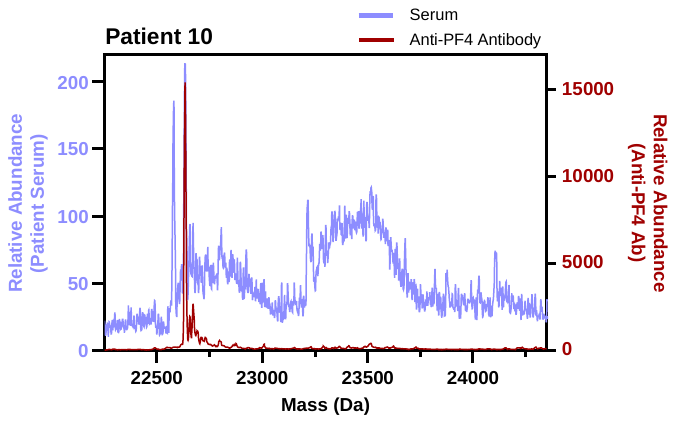

| A. 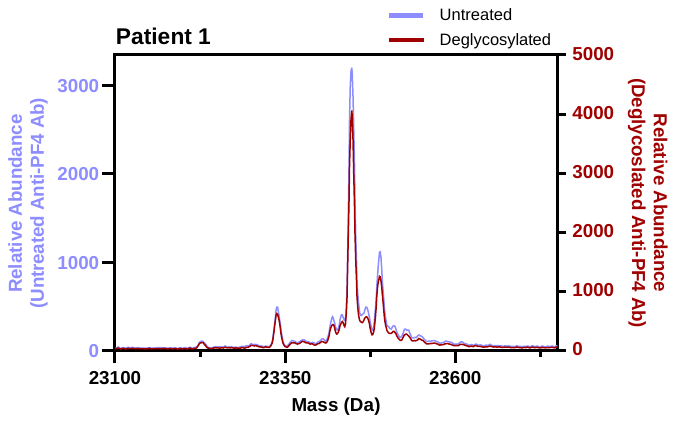 | D. 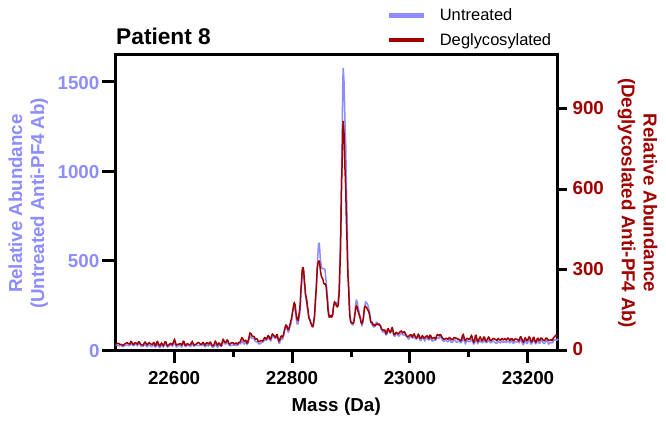 |
| --- | --- |
| B. 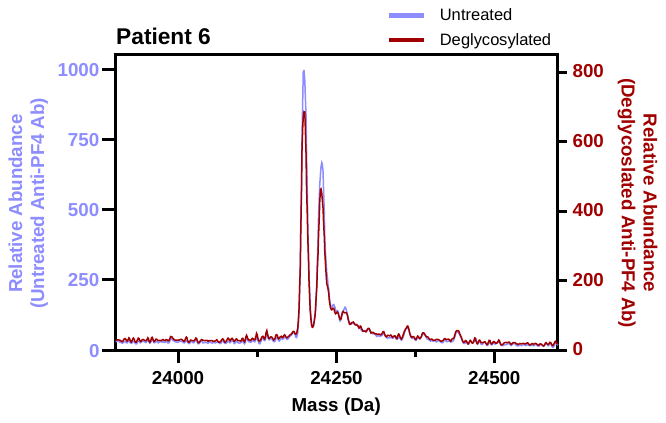 | E. 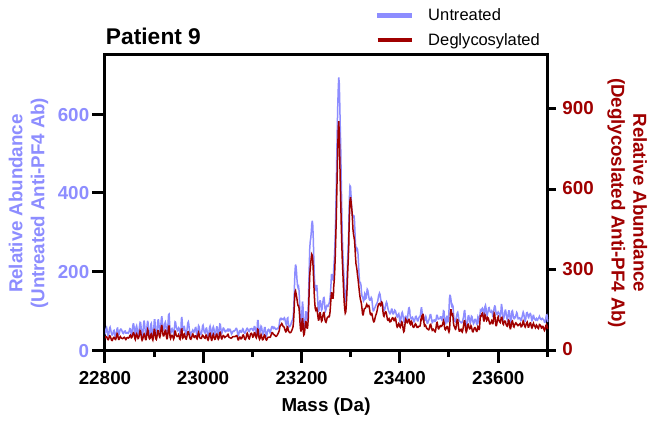 |
| C. 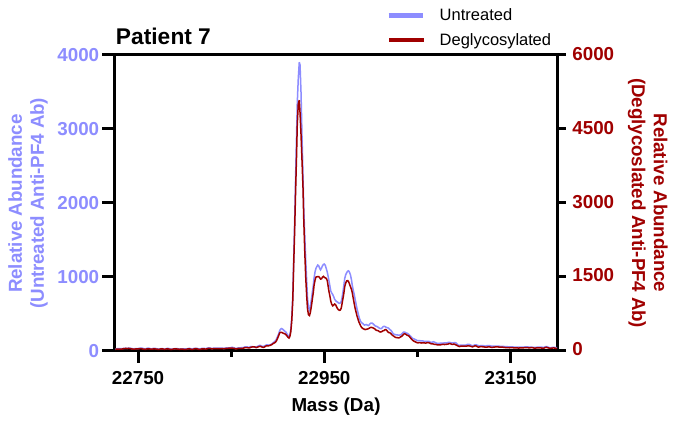 | F. 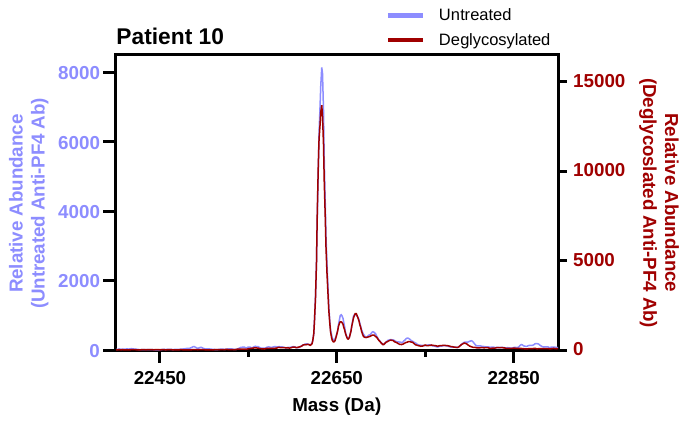 |

**Figure S3.** **Deglycosylation of isolated anti-PF4 antibodies demonstrates no change in light chain profiles. (A-F)** LC-ESI-QTOF calculated mass spectra (Daltons) displaying light chain distributions associated with IgG heavy chain of untreated (blue) and PNGase digested (maroon) isolated anti-PF4 antibodies. The y-axis presents the relative abundance of identified antibodies.

**Methods**

*Patient Samples*

Patient serum samples were obtained from fifteen individuals diagnosed with HIT. Research studies were approved by the Institutional Review Board of Mayo Clinic.

*ELISA Studies*

PF4 IgG (Immucor) testing was performed as ordered by the treating physician. Zymutest HIA IgG (Hyphen BioMed) immunoassay was performed according to manufacturer instructions. In brief, patient samples were incubated with platelet lysate coated plates. The plates were washed then incubated with an anti-human IgG (Fc-specific) peroxidase antibody. After antibody incubation, tetramethylbenzidine in presence of hydrogen peroxide was added. The reaction was stopped with sulfuric acid, and colorimetric detection was performed at optical density 450 nm.

*Functional platelet studies*

Serotonin release assay (SRA) testing was performed as ordered by the treating physician. The PF4-dependent P-selectin expression assay (PEA) was performed as previously described.^1,2^ Prostaglandin E1 was added to citrated whole blood obtained from healthy volunteers to a 50 ng/mL final concentration. Whole blood was centrifuged at 200 x g for 15 minutes to obtain platelet-rich plasma. Platelet isolation was performed by centrifugation at 1,000 x g for 10 minutes followed by resuspension of the platelet pellet in PBS/1.0% BSA. Platelets (1 x 10^6^) were then treated for 20 minutes at ambient temperature with PF4 (37.5 μg/mL) or PBS. 10 μL of the patient sample and 40 μL of platelets were incubated for 1 hour at ambient temperature before the addition of fluorescently labeled anti-P-selectin (monoclonal antibody HB-299, ATCC) and anti-GPIIIa (monoclonal antibody HB-242, ATCC) antibodies for 20 minutes. A final volume of 200 µL was then obtained by the addition of PBS-BSA. Platelet events were gated based on GPIIIa positivity before P-selectin expression (median fluorescence intensity, MFI) was recorded.

*Antibody Isolation*

Anti-PF4 antibodies were isolated as described^3,4^. Briefly, heparin Sepharose beads (200 μL, Cytiva Lifesciences) were washed with PBS, pH 7.4, and incubated with 200 μg of recombinant PF4 for one hour. Beads were then blocked with PBS/0.1% BSA for 30 minutes. Following a thorough washing with PBS 250 μL of patient samples were added to beads for 1 hour. Beads were thoroughly washed with PBS, and eluates were obtained from the PF4/heparin Sepharose beads using 2M NaCl. Eluates were dialyzed against PBS before being evaluated by ELISA, PEA, and mass spectrometric studies.

*Deglycosylation of antibodies*

Antibody deglycosylation was performed per manufacturer instructions (NEB). Briefly, 125 μL of anti-PF4 antibody eluate and 1 μl of PNGase F were added to the reaction mixture and mixed gently. The reaction was then incubated at 37 C for 24 hours.

*Matrix-assisted laser desorption ionization time-of-flight Mass Spectrometry*

Mass-Fix was performed on patient samples as previously described^5^. CaptureSelect resins (Thermo Fisher) were washed with 30 volumes of PBS + 0.1% Tween-20. Using a final working volume of resins of 10% w/v, 50 μL of each CaptureSelect Resin solution was pipetted into the wells of a 384-well plate, and 10 μL of patient serum were added to each well. After incubating for 15 minutes, the serum supernatant was removed from the resin. The resin was washed using 50 μL of PBS 3 times and 50 μL of water 3 times. Captured Igs were then eluted and reduced using 30 mcl of 20 mM TCEP + 0.10% TFA. Finally, sample eluates were diluted 1:2 in a separate dilution plate, using TCEP + 0.10% TFA. A ttpLabtech Mosquito nanoliter pipettor (Hertfordshire, United Kingdom) was utilized for spotting. Sample eluate (0.5 μL of purified patient Igs in 0.1% TFA containing 10 mM TCEP) was combined with alpha-cyano-4-hydroxycinnamic acid (CHCA) matrix (0.7 μL, 10 mg/mL in 50% ACN + 0.1% TFA) and spotted onto a 96-well microScout polished steel Bruker target (Bruker Daltonics) using a single application spotting method. Analysis was performed in positive ion mode with a summation of 500 laser shots using a MALDI-TOF mass spectrometer (Bruker Microflex LT, Germany).

*Liquid Chromatography Electrospray Ionization time-of-flight mass spectrometry*

The basic method used for antibody analysis has been previously described.^6,7^ Immunoglobulins (Igs) from patient sera or bead eluates were isolated using camelid-derived nanobodies selective for the constant domains of human Ig gamma heavy chain, kappa light chain, or lambda light chains (CaptureSelect affinity resins, Thermo Fisher Scientific). 100 μL or 50 μL of camelid nanobody beads were incubated with 10 μL of serum or 100 μL of anti-PF4 antibody eluate, respectively, and incubated for 30 minutes at ambient temperature. Subsequently, sera or eluate supernatants were removed, and the beads were washed 3 times with 500 μL of water. Ig from light and heavy chains of sera and anti-PF4 antibody eluate were eluted using 60 μL or 20 μL of 5% acetic acid, respectively. Following a 5-minute incubation, eluted Igs were reduced using 100 mM dithiothreitol in 1M ammonium bicarbonate (2:1; v:v) to separate light chain and heavy Ig chains. An Agilent 1290 Infinity II liquid chromatography (LC) system was used to separate Ig chains before ionization and to remove any co-eluted PF4 before analysis using a SCIEX Zeno time-of-flight (TOF) 7600 mass spectrometer (MS). 10 μL of each camelid nanobody bead eluate were injected per analysis onto a Poroshell 300SB-C3 column (2.1 mm X 75 mm) with a 5 μm particle size placed in a 60 ºC column heater. The mobile phases included an aqueous phase A (100% water + 1% formic acid) and an organic phase B (90% acetonitrile + 10% isopropanol + 0.1% formic acid), and the flow rate was 300 μL/min. The light chains elution range was identified during a 4.5-minute gradient from 27%B to 32%B. The diverter valve was used to direct 10.35 minutes of the gradient into the MS; otherwise, the LC was diverted to waste. The MS, using positive electrospray ionization, was run using intact protein workflow; CUR 30, CAD 7 GAS1 35, GAS2 30, and source temperature 500 °C. TOF MS data from collected from 600 to 2500 m/z; DP 175 and CE 10. Data analysis was performed using Sciex OS v2.2 and PeakView ver. 2.2. Mass spectra were analyzed using the [M + 12H]^12+^ light chain charge state to detect clones as described elsewhere.^6,7^ The retention time of the monoclonal light chain in each patient sample was tracked using PeakView. The mass spectra of the multiply charged light chain ions were deconvoluted to obtain an accurate molecular mass using the Bio Tool Kit ver. 2.2 plug-in software.
